# Forecasting the spread of COVID-19 pandemic in Bangladesh using ARIMA model

**DOI:** 10.1101/2020.10.22.20217414

**Authors:** Lakshmi Rani Kundu, Most. Zannatul Ferdous, Ummay Soumayia Islam, Marjia Sultana

**Affiliations:** Department of Public Health and Informatics, Jahangirnagar University, Savar, Dhaka-1342, Bangladesh; Department of Computer Science and Engineering, Begum Rokeya University, Rangpur-5400, Bangladesh

**Keywords:** COVID-19, Confirmed cases, Deaths, Forecast, ARIMA, Bangladesh

## Abstract

**Background:** COVID-19 is one of the most serious global public health threats creating an alarming situation. Therefore, there is an urgent need for investigating and predicting COVID-19 incidence to control its spread more effectively. This study aim to forecast the expected number of daily total confirmed cases, total confirmed new cases, total deaths and total new deaths of COVID-19 in Bangladesh for next 30 days.

**Methods:** The number of daily total confirmed cases, total confirmed new cases, total deaths and total new deaths of COVID-19 from 8 March 2020 to 16 October, 2020 was collected to fit an Autoregressive Integrated Moving Average (ARIMA) model to forecast the spread of COVID-19 in Bangladesh from 17^th^ October 2020 to 15^th^ November 2020. All statistical analyses were conducted using R-3.6.3 software with a significant level of *p< 0*.*05*.

**Results:** The ARIMA (0,2,1) and ARIMA (0,1,1) model was adopted for forecasting the number of daily total confirmed cases, total deaths and total confirmed new cases, new deaths of COVID-19, respectively. The results showed that an upward trend for the total confirmed cases and total deaths, while total confirmed new cases and total new death, will become stable in the next 30 days if prevention measures are strictly followed to limit the spread of COVID-19.

**Conclusions:** The forecasting results of COVID-19 will not be dreadful for upcoming month in Bangladesh. However, the government and health authorities should take new approaches and keep strong monitoring of the existing strategies to control the further spread of this pandemic.

## Introduction

The COVID-19 pandemic is an ongoing public health threat which is caused by severe acute respiratory syndrome coronavirus 2 (SARS-CoV-2) ^1^. It is a viral infection and highly infectious disease considering to be transmitted from wild animals (bats) to human and first identified in Wuhan city of China ^2,3^. The disease was then a local epidemic of China, but soon it became expanded all over the world by international travelers ^4^. Previously SARS-CoV emerged from China in 2003, respectively MERS-CoV emerged from Middle East in 2012, developed severe symptoms ^5,6^. Now this virus is the 7^th^ of coronaviruses that represents a serious public health threat for which World Health Organization (WHO) declared COVID-19 as a pandemic in March 2020 ^7^. Since then, the pandemic has spread all over the globe as days go by. Approximately, 190 countries have been affected, where major outbreaks occurred in USA, Italy, Spain, France, China ^4^. As of 14^th^ October, 2020, COVID-19 cases have been exceeded 38 million including 1,083,234 deaths worldwide ^8^. According to a study conducted in China, COVID-19 can be asymptomatic including mild or moderate symptoms ^9^. Hence, by looking into these facts, we should have to imagine the heaviness of this pandemic globally and its impacts on public health.

In Bangladesh, Institute of Epidemiology, Disease Control and Research (IEDCR), declared the first three confirmed cases of COVID-19 on 8^th^ March, 2020 ^10^. The present scenario of coronavirus cases in Bangladesh is 3,91586 confirmed cases, with 5699 deaths, and 3,07141 recoveries on 20 October 2020 ^11^. As Bangladesh is one of the most densely populated countries around the world, it has a great risk of exposure due to COVID-19. Like other countries, government of Bangladesh has also already adopted several measures such as informing COVID-19 hotspot areas, maintaining social distance and increasing mass awareness by social media or televisions, setting lockdown of school, college and office ^12^ to minimize the situation. However, these available control measures are significantly influenced by the knowledge, attitudes, and practices (KAP) towards COVID-19 ^13^. Furthermore, it is really a challenging task for the people of overcrowded Bangladesh where the chance of the COVID-19 spreading is much more than non crowded place and in a Bangladesh survey from March 29 to April 29 found that 98.7% reported wearing a face mask in crowded places ^13^. As the incubation period of COVID-19 is up to 14 days, the virus can be transmitted to other people during this time period ^14,15^. Again, there is ambiguity about the proper decline and fall of the contagious disease ^16^.

Therefore, in this critical time, smart planning with sufficient preparation for mitigating the incidence and prevalence of disease including designing the future prospect is very important. Because evidence on the management approaches of current COVID-19 pandemic is still limited though the numbers of affected countries are increasing as the days go by ^17^. By modeling a future forecast which estimates the regular number of confirmed cases might help to implement new rules. Further, a statistical forecast model might also be beneficial for predicting future epidemic threat as well as better management of societal, economic, cultural and public health matters ^18,19^.

The aim of this study is to predict the spread and the final size of COVID-19 epidemic in Bangladesh by using Auto Regressive Integrated Moving Average (ARIMA) model. The ARIMA model is generally known as Box-Jenkins methodology used to forecast and analysis in a time series modeling approach ^20^. Recently this model has been used in the mostly affected 15 countries of the world to forecast the flow of COVID-19 which revealed similar number according to the current situation of those countries ^21^. Another study conducted in Italy and Spain showed accurate regular number of cases in these countries ^22^. Though modeling cannot always forecast the accurate number of cases it may help to summarize the future prospect of the pandemic by showing the acceptable number of occurrence happening for the next 30 days.

## Methods and materials

### Data Source and data description

The data extracted from the official website of the World Health Organization COVID-19 situation reports ^23^ and website of the Humanitarian Data Exchange (https://data.humdata.org/dataset/coronavirus-COVID-19-cases-and-death).

This study considered the daily total confirmed cases, daily total confirmed new cases, total deaths and total new deaths of COVID-19 from 8^th^ March (day 1) to 16^th^ October 2020 (day 223) in Bangladesh. Then, this data were used to fit a best ARIMA model for forecasting next 30 days total confirmed cases or new cases as well as total deaths or new deaths from 17^th^ October (day 224) to 15^th^ November (day 253) of COVID-19 in Bangladesh.

### Statistical Analysis

All analyses were done with R software (version 3.6.2), the stationarity check was conducted using ‘tseries’ package and ARIMA model was fitted using ‘forecast’ package. A *p*-value of less than 0.05 was considered statistically significant.

### ARIMA Model description

In this study, a linear parametric Autoregressive Integrated Moving Average (ARIMA) model was applied for prediction purpose. If a time series *Y*_*t*_ is followed by *Y*_*t*_ = *θ*+ *α*_1_*Y*_*t*−1_ + *α*_2_*Y*_*t*−2_ + … + *α* _*p*_*Y*_*t*− *p*_ + *u*_*t*_, where *u*_*t*_ is a white noise with mean zero and variance *σ* ^*2*^, then it is called an autoregressive process of order *p* and is denoted by *AR(p)*. If *Y*_*t*_ is defined by *Y*_*t*_ = *β*_1_*u*_*t*−1_ + *β*_2_*u*_*t*−2_ + … + *β* _*p*_*u*_*t*−*q*_, then it is called a moving average process of order *q* and is denoted by *MA(q)*.

The combination of AR and MA models are known as ARMA model. An *ARMA(p,q)* model is given by *Y*_*t*_ = *α*_1_*Y*_*t* −1_ + *α*_2_*Y*_*t* −2_ + … + *α* _*p*_*Y*_*t* − *p*_ + *u*_*t*_ + *β*_1_*u*_*t* −1_ + *β*_2_*u*_*t* −2_ + … + *β* _*p*_*u*_*t* −*q*_ ^24^. A time series *Y*_*t*_ is said to follow an Autoregressive Integrated Moving Average (ARIMA) model if the *d^th^* difference *W* = ∇^*d*^*Y_t_* is a stationary ARMA process. If *W*_*t*_ follows an *ARMA(p,q)* model, then we say that *Y*_*t*_ is an *ARIMA(p,d,q)* process. For practical purposes, taking d=1 or at most 2 ^25^. Thus, an *ARIMA (p, 1, q)* process with *W*_*t*_ = *Y*_*t*_ −*Y*_*t*−1_ can be written as

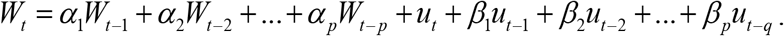

### Box-Jenkins Method

ARIMA model also known as Box-Jenkins method which is widely used for the analysis of time seriesand forecasting ^26^. In recent years, this model has been widely used in the prediction of epidemic trend of infectious diseases ^27^. The fitting of the ARIMA model or Box-Jenkins methodology consists of the following steps:

### Test of Stationarity

An Augmented Dickey-Fuller (ADF) test wasconducted for testing whether the original series is stationary or not ^28^. Difference or logarithmic transformation was adapted to transformed nonstationary time series into a stationary time series. Achieving stationary is a precondition for establishing an ARIMA model.

### Model Identification

An appropriate value of p, d and q of ARIMA model was identified. The value of d was identified according to the number of differentials. From the autocorrelation function (ACF) and partial autocorrelation function (PACF) plot against the lag length, the AR and MA parameters was selected. Nevertheless, there have been some model selection criteria such as Akaike information criterion (AIC), Bayesian information criteria (BIC). The optimal model was chosen based on smallest value of AIC and BIC ^29^.

### Estimation of the fitted Model

After the identification of the appropriate values of p and q, the next stage was to estimate the parameter of the autoregressive and moving average terms included in the model. This was done using the maximum likelihood estimation method.

### Diagnosting Checking

Having identified the fitted ARIMA model and parameter estimate, the Ljung-Box Q test was applied to check whether the residual series is a white noise. If so, then the fitted model was accepted. The Ljung-Box Q statistics is defined as

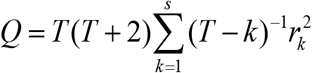

where, T is the number of observations, s is length of coefficients to test autocorrelation, *r*_*k*_ is autocorrelation coefficient for lag k. This statistic *Q* approximately follows the chi-square distribution with *(k-q)* degrees of freedom, where *q* is the number of parameter should be estimated in the model ^30^.

### Forecasting

Finally, the future value was predicted by using the fitted model. The steps are presented in the following diagram:

### Ethics

As all data were obtained from secondary data collection source, no formal ethical assessment was required.

## Results

The time series plot of daily COVID-19 total confirmed new cases (A1), total confirmed cases (A2), total new deaths (B1) and total deaths (B2) in Bangladesh from March 8 to October 16, 2020 are presented in figure 2. During the study period, a total of 384,559 confirmed cases and 5,608 deaths were detected and there were maximum cases in 3^rd^ July 2020 of 4019 cases and maximum deaths in 1^st^ July 2020 of 64 deaths. The graphical inspection showed that the original series are in increasing trend and sometimes are in decreasing trend and the variance is not stable which leads the variables were nonstationary and need to be transformed into a stationary process.

**Figure 1:**
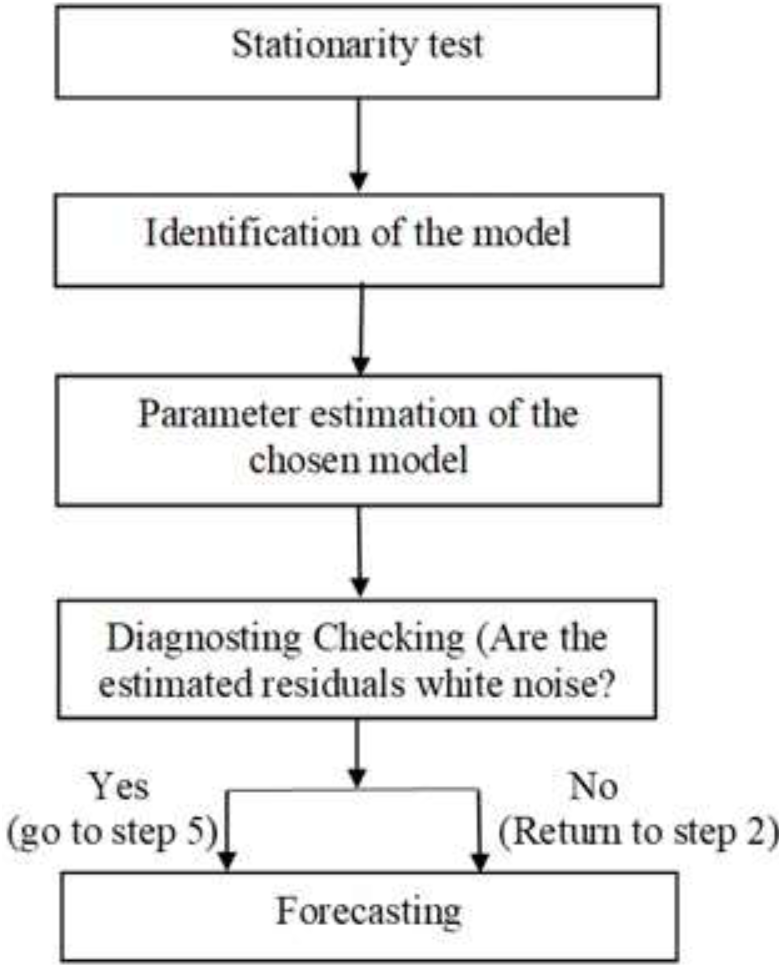
Scheme for the use of Box-Jenkins methodology^31^

**Figure 2.**
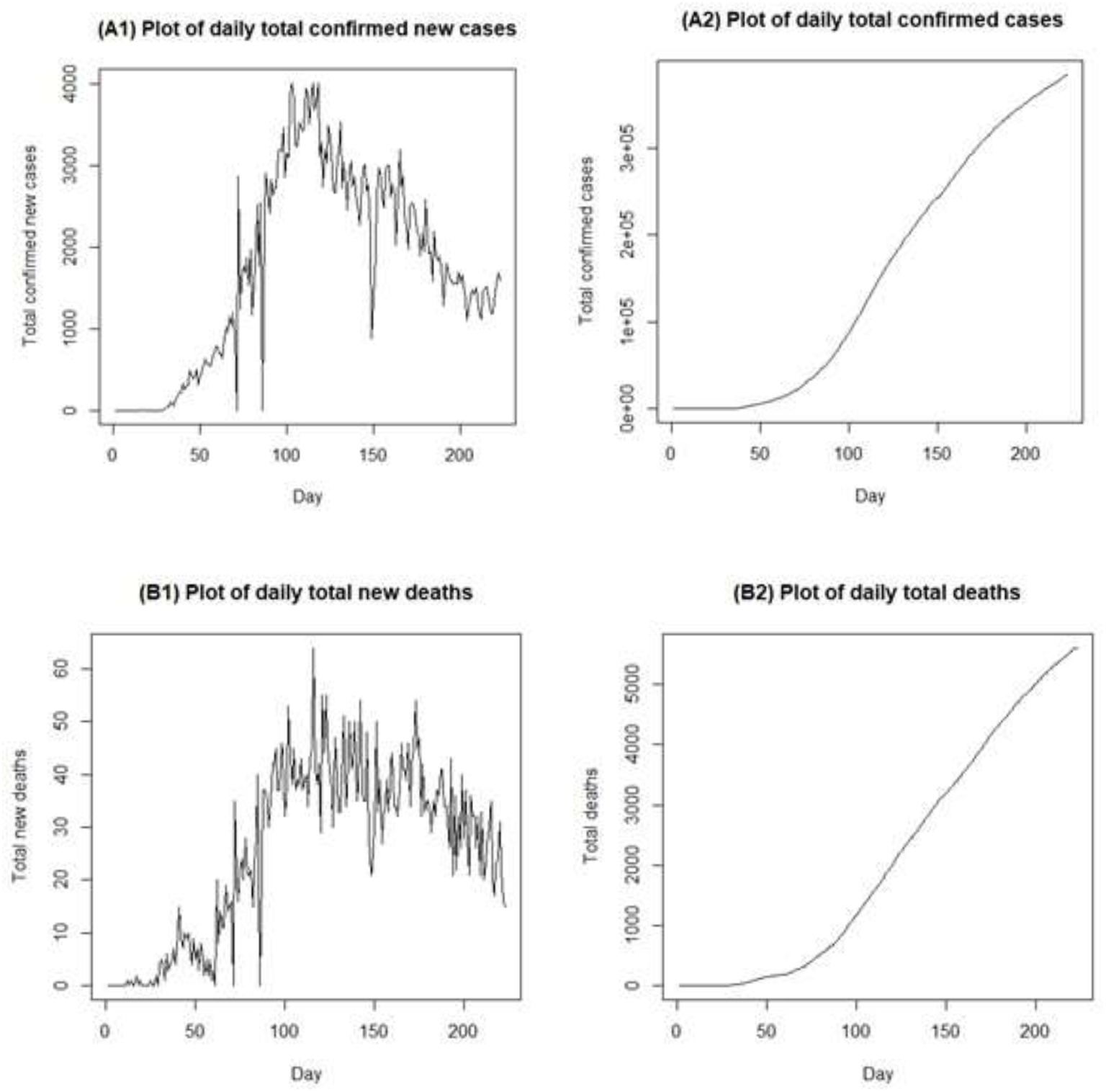
Time series plot displays for confirmed new cases (A1), total confirmed cases (A2), new deaths (B1) and total deaths (B2) of COVID-19 in Bangladesh.

Before fitting the ARIMA model, it is necessary to confirmed that the series must be stationary. Augmented Dickey Fuller (ADF) test was applied to check the stationarity of the series. The ADF unit root test results are presented in Table 1.

**Table 1.** Results of Augmented Dickey Fuller unit root test

| Variables | Original data |  | At 1<sup>st</sup> difference |  | At 2<sup>nd</sup> difference |  |
| --- | --- | --- | --- | --- | --- | --- |
|  | ADF | *p*-value | ADF | *p*-value | ADF | *p*-value |
| Total confirmed new cases | -0.4148 | 0.985 | -8.135 | 0.01* | - | - |
| Total new Deaths | -0.280 | 0.9831 | -7.709 | 0.01* | - | - |
| Total confirmed cases | -3.609 | 0.063 | -0.429 | 0.984 | -8.122 | 0.01* |
| Total Deaths | -2.775 | 0.25 | -0.290 | 0.99 | -7.698 | 0.01* |

The null hypothesis is that the series is non-stationary, or contain a unit root. Decision Rule: Reject the null hypothesis if the $p\text{-value} < \alpha = 0.05$ ; \* indicates the rejection of the null hypothesis.

The findings indicated that all the series are nonstationary at their level (p-value>0.05), but after taking 1^st^ difference, total confirmed new cases and deaths achieved stable variance. On the other hand, total confirmed cases and deaths achieved stable variance after 2^nd^ differences. This ensured that the series are stationary at 5% level of significance and ready for the Box-Jenkins ARIMA modeling approach.

After achieving the stationary series, a list of potential models was formulated based on the significant spikes observed from the autocorrelation function (ACF) and partial autocorrelation function (PACF). Among the candidate models ARIMA(0,2,1) model produces the lowest AIC, AICc, BIC for the total confirmed cases and total deaths and ARIMA(0,1,1) model exhibits the lowest AIC, AICc, BIC for confirmed new cases and new deaths which are demonstrated in Table 2.

**Table 2:**
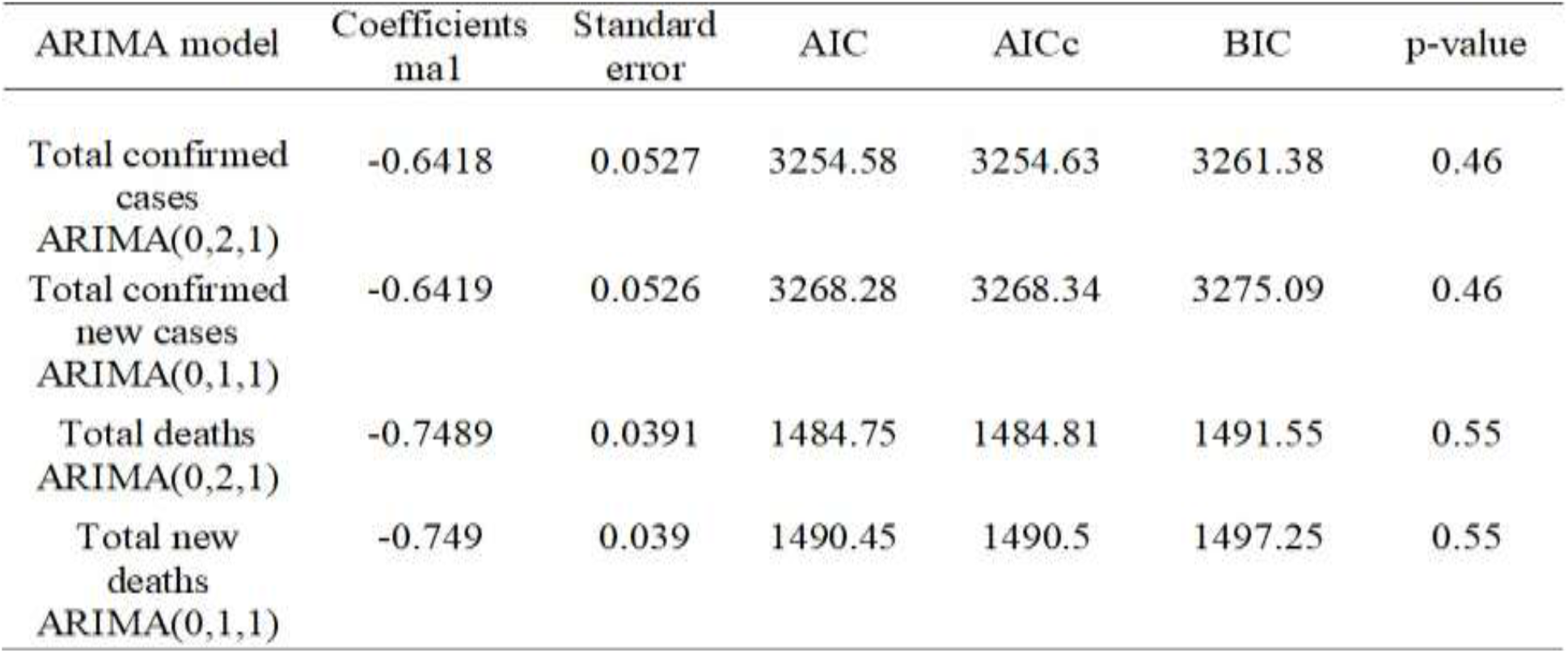
Results of each selected ARIMA model and Ljung-Box test

The Ljung-Box Q test suggested that the residuals series of the ARIMA (0,2,1) and ARIMA (0,1,1) model are purely white noise (*p*-value>0.05) at 95% confidence level. Therefore, these selected models are probably adequate for the data. Then these models were applied to forecast the daily confirmed cases and deaths of COVID-19 in Bangladesh.

Table 3 and figure 3 showed the predicted values from 17^th^ October to 15^th^ November 2020 for all variables using the fitted ARIMA(0,2,1) and ARIMA(0,1,1) model with 95% confidence interval (CI). The forecasted value (in blue) based on fitted ARIMA model for daily confirmed cases or new cases and deaths or new deaths of COVID-19 for the next 30 days and the current number of confirmed cases and deaths from March 8, 2020 to October 16, 2020 (in black) are shown in figure 3. The results showed that an upward trend for daily total confirmed cases and total deaths in Bangladesh by 15^th^ November, 2020 has a point forecast of 430828 (95% CI 402683-458974) and 6228 (95% CI 5849-6608), while total confirmed new cases and total new death, possible become stable. However, Bangladesh is hopeful to control this pandemic at the middle of November.

**Table 3:**
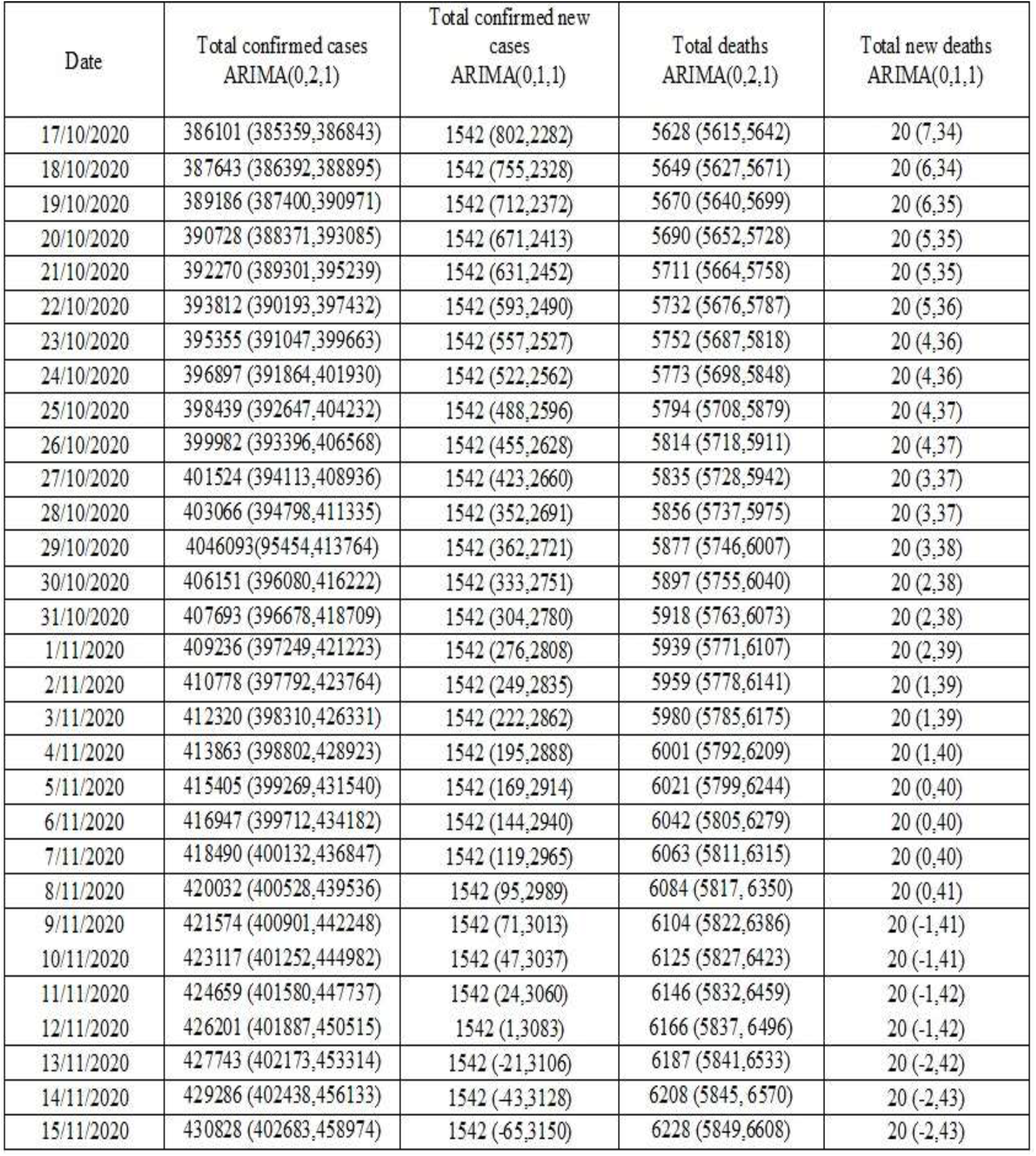
Forecasting of daily total confirmed cases, total confirmed new cases, total deaths, and total new deaths in Bangladesh for the next 30 days according to ARIMA models with 95% CI

**Figure 3:**
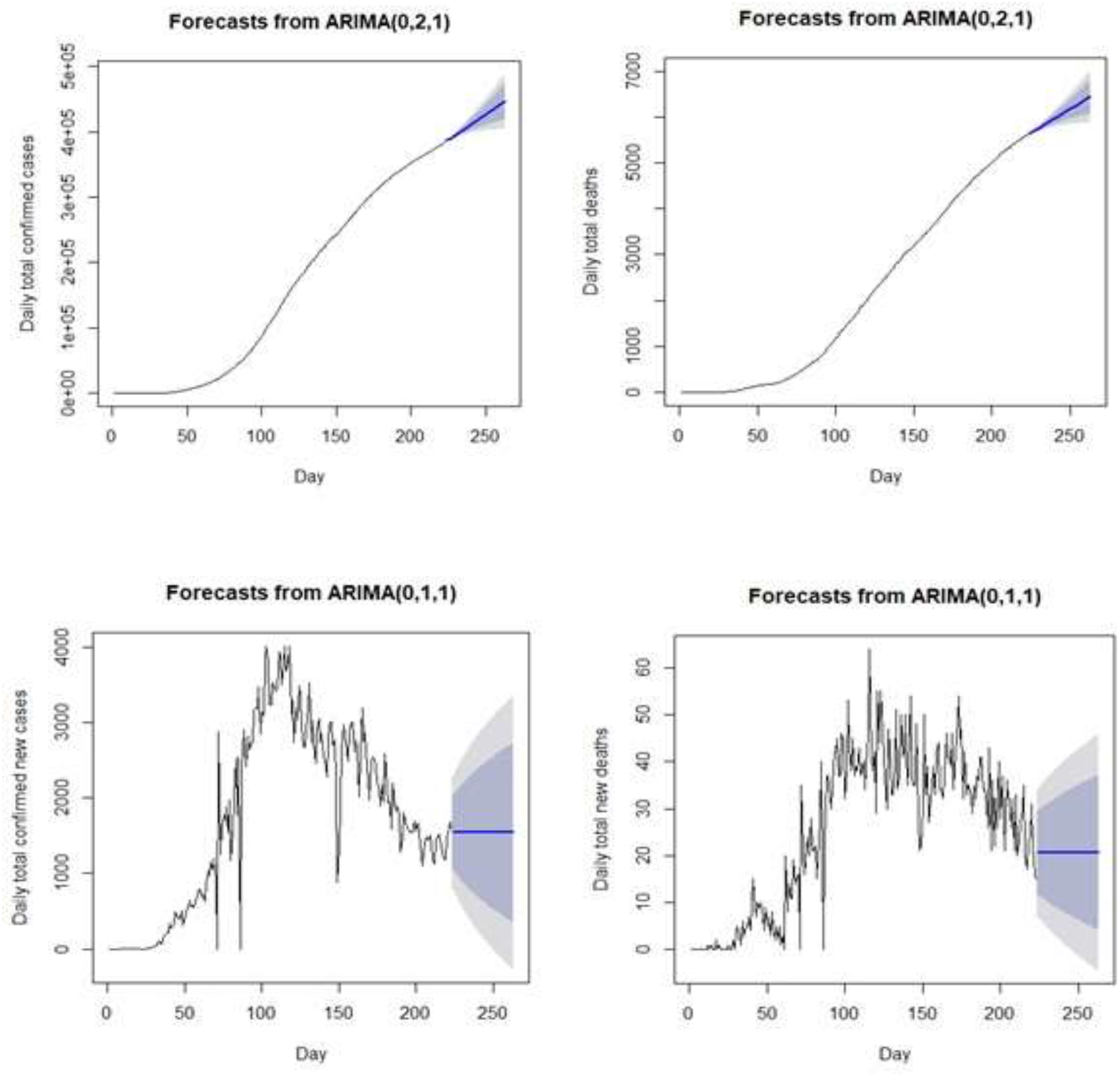
Predictive and confidence intervals of daily total confirmed cases, total confirmed new cases, total deaths, total new deaths of COVID-19 using fitted model (Black line: actual data, Blue line: 30 days forecast, Gray zone: 80% of CI, White zone: 95% of CI).

## Discussion

It is alarming that the number of novel coronavirus (SARS-Cov-2) cases continues to escalate across the globe. The estimation of infectious disease like COVID-19 management by developing hypothesis for interpreting the observed situation can be done via time series analysis ^32^.Time series health researchers widely use ARIMA model because of the importance of ‘time’ for disease management studies ^33,34^. Previous study revealed that ARIMA is one of the most suitable models as it has higher fitting and forecasting accuracy ^35,36^. In recent years, it is also an useful model for predicting the incidence of infectious disease ^20^. The main purpose of this work is to monitor and forecast the expected number of the new COVID-2019 patients in Bangladesh by applying a commonly used time series model, known as an ARIMA model, based on the data of the total confirmed daily cases and deaths and the new confirmed cases and new deaths officially announced by the Institute of Epidemiology, Disease Control and Research.

The prediction indicates an upward trend for daily total confirmed cases along with total deaths. At the same time, total confirmed new cases, and total new deaths most probably become stable. According to the study the total confirmed cases of coronavirus in 18^th^ October, 2020 was estimated 388,569 and total death was estimated 5660 which was nearly similar to the actual scenario^37^. The present reveals that, Bangladesh is hopeful for controlling the pandemic at the mid of November 2020 if the spreading pattern of the disease remains the same. A study conducted in Iran reported that, an upward trend for total confirmed case and total death while the other variables such as total confirmed new cases, total new deaths possibly became steady which is similar to our study ^38^. Another study conducted for forecasting different countries COVID-19 trend demonstrated stable condition for China, stationary trend for South Korea while Thailand showed controlled condition ^18^. Based on a study, for predicting the end of COVID-19 by using this model expected that top countries COVID-19 infection will slow down by October, 2020 ^39^. In contrast, in Saudi Arabia and Nigeria this model forecasts highly increased of daily case with cumulative daily cases within one month ^40,41^. Similarly another study conducted India revealed explicit rising of infection in coming days especially in west and south Indian regions are more at risk ^42,43^.

Although, ARIMA model usually gives better forecast, few drawbacks exits. Firstly, it does not have automatic updates. Secondly, if more data added in the study then model gives different forecast results. Thirdly, the prediction accuracy has a direct relation with the number of observation.

## Conclusion

In this study, the trend of COVID-19 outbreak in Bangladesh was observed. The results found that the best prediction model is ARIMA (0,1,1) for forecasting the trend of the number of daily new confirmed cases and deaths in Bangladesh. In this model the number of daily confirmed cases and deaths showed upward trend. But surprisingly model presents the number of new confirmed cases and new deaths will become stable in next 30 days. It is hopeful for Bangladesh, a developing country, though having inadequate medical facilities, becoming successful to control this pandemic within last few days. However, to prevent the COVID-19 pandemic permanently until proper vaccine or medicine is developed, public health authorities, government, and non-government should take hard decisions to control the further increase of this pandemic. Besides all the authorities, the general public should maintain social distance and undertake all necessary preventive measures to stay free from the disease and control its spread.

## Data Availability

The data underlying this article were accessed from World Health Organization COVID-19 situation reports and website of the Humanitarian Data Exchange. The corresponding author has no right to share the data.

